# Validation study for noninvasive single-cell-based prenatal genetic testing

**DOI:** 10.1101/2023.08.29.23294301

**Authors:** Michelle Bellair, Elisabete Amaral, Mason Ouren, Cameron Roark, Jaeweon Kim, April O’Connor, Adrianna Soriano, Margaret L. Schindler, Ronald J. Wapner, Joanne L. Stone, Nicola Tavella, Audrey Merriam, Lauren Perley, Amy M. Breman, Arthur L. Beaudet

## Abstract

**Objective:** To clinically validate a cell-based noninvasive prenatal genetic test using sequence-based copy number analysis of single trophoblasts from maternal blood.

**Methods:** Blood was obtained from 401 individuals (8-22 weeks) and shipped overnight. Red blood cells were lysed, and nucleated cells stained for cytokeratin (CK) and CD45 using fluorescent antibodies and enriched for positive CK staining. Automated microscopic scanning was used to identify and pick single CK^+^/CD45^-^ trophoblasts which were subjected to whole genome amplification and next-generation sequencing.

**Results:** Blood was obtained from 243 pregnancies scheduled for CVS or amniocentesis. Luna results were normal for 160 singletons while 15 cases were abnormal (14 aneuploidy and one monozygotic twin case with Williams syndrome deletion). These Luna results agreed with CVS/amniocentesis. Placental mosaicism occurred in 7 of 236 (3.0%) Luna cases and in 3 of 188 (1.6%) CVS cases (total 4.6%). No scorable trophoblasts were recovered in 32 of 236 (13.6%) usable samples. Additionally, 158 low-risk pregnancies not undergoing CVS/amniocentesis showed normal results for 133 cases. Seven had aneuploidy results, and there were 3 likely pathogenic deletions or duplications including one15q11-q13 deletion.

**Conclusion:** This noninvasive cell-based prenatal genetic test detected aneuploidy and deletions/duplications with high sensitivity and specificity based on concordance with CVS/amniocentesis.

**Key points:** *What’s already known about this topic?:*

- As a proof of principle for noninvasive genetic prenatal diagnosis, circulating fetal trophoblasts have been isolated from maternal blood and analyzed for detection of aneuploidy and genomic deletions and duplications.
- These trophoblasts reflect the genotype of the current placenta(s) but not necessarily the genotype of the fetus because of placental mosaicism.

*What does this study add?:*

- This study demonstrates the advantages of single cell analysis and the feasibility of launching a test for reliable detection of cytogenetic aneuploidy, deletions, and duplications.
- This test has improved detection of deletions and duplications compared to cell-free NIPT, but widespread adoption will require improved recovery of fetal cells from maternal blood and reduced cost through automation and high-throughput.

## 1 INTRODUCTION

Currently, the most widely used laboratory procedures for prenatal genetic testing are amniocentesis, chorionic villus sampling (CVS), and cell-free noninvasive prenatal testing (cfNIPT). Tests performed on CVS and amniocentesis specimens are considered diagnostic but are invasive and carry a small risk of pregnancy loss.^1^ In contrast, cfNIPT is a risk-free screening test, but the ability to detect chromosomal deletions and duplications is inferior to what is achievable with CVS and amniocentesis. The cfNIPT also produces varying frequencies of false positive results, depending on the nature of the abnormality detected and the genomic region involved. The availability of a prenatal genetic test that is risk-free and provides high resolution detection of pathogenic deletions and duplications would be a superior option for patients and clinicians alike.

The potential to use circulating fetal cells in maternal blood to derive noninvasive prenatal genetic results has been hypothesized since at least 1969,^2^ but various technological challenges, primarily the extreme rarity of fetal cells in maternal circulation, have hindered the introduction of such a test into the clinic. Although there is evidence that fetal nucleated red blood cells (fnRBC), fetal lymphocytes, and fetal hematopoietic stem progenitor cells (HSPC) can be recovered from maternal blood, none of these have achieved full clinical utility. Steen Kølvraa and colleagues presented evidence in 2014 that extravillous trophoblasts (EVTs) might be a suitable target for cell-based noninvasive prenatal testing.^3,4^ Using CD105 and CD141 as a surface markers and cytokeratin as a cytoplasmic marker for these trophoblasts, they detected fetal cells in 91% of 78 samples which was confirmed by XY fluorescence in situ hybridization (FISH). A next key development in 2011 when it was demonstrated that DNA from single human tumor cells could be used for next-generation sequencing (NGS) that would be clinically impactful.^5^ Furthermore, it was shown in 2012 that clinically significant deletions and duplications could be detected in single human lymphoblasts at a resolution of 1 Mb using array comparative genomic hybridization (CGH).^6^ In 2016, two collaborative publications reported further progress in using isolated circulating trophoblasts to detect fetal aneuploidy and deletions.^7,8^ This proof of feasibility for prenatal diagnosis was further confirmed using NanoVelcro microchips.^9^ Results from genomic copy number analysis of trophoblasts from maternal blood compared favorably with those from CVS or amniocentesis.^10^ Most of these reports focused on detection of aneuploidy, deletions, and duplications, but the ability to detect monogenic disorders has also been reported.^11,12^ Most recently, two groups have reported continuing success performing cell-based NIPT.^13,14^ Multiple reviews from different eras are available.^15–17^ We report a series of clinical validation studies designed to demonstrate the feasibility of a clinical high-resolution noninvasive single-cell-based prenatal test using whole genome NGS for detection of aneuploidy and deletion/duplication.

## 2 METHODS

### 2.1 Study Design

Three studies (Table 1, insert table) were carried out for validation of the cell-based noninvasive Luna Prenatal Test (hereafter referred to as the Luna test). These included a **no-follow-up (NFU) series** obtained from routine pregnancies, a **follow-up (FU) series** scheduled to have CVS or amniocentesis, and a **spike-in series** of cells from seven lymphoblast or fibroblast cell lines with known aneuploidy or deletion/duplication (del/dup). The **NFU series** was designed to assess the success rate for recovering trophoblast cells and for achieving NGS analyses that could be evaluated for either aneuploidy only, or for aneuploidy plus del/dup at a resolution of 1.5 Mb for deletions and 2 Mb for duplications. Blood was collected between Jul. 2021 through Jan. 2022 from healthy pregnant volunteers who were distributed across 35 states in the USA. Participants were recruited through social media and consented virtually by a genetic counselor employed by Luna. A phlebotomist visited the home, drew blood, and delivered the sample for shipping. Each sample was processed with the methods that were later used for launch of the Luna clinical test. About a third of the way through the study, an initiative to collect a second blood sample was implemented for samples where 0 or 1 cells were recovered in the first sample. An overview of sample characteristics in the **NFU series** is provided in Table 2, insert table.

**TABLE 1.** Three groups of validation samples.

| **Study** | **Sample number** | **Characteristics** |
| --- | --- | --- |
| No-follow-up (NFU) | 158 cases | From routine pregnancies. No CVS or amnio |
| Follow-up (FU) | 243 cases | Scheduled for CVS or amnio |
| Spike in | 148 cells | Seven cell lines with aneuploidy, deletion, or duplication |

**TABLE 2.**
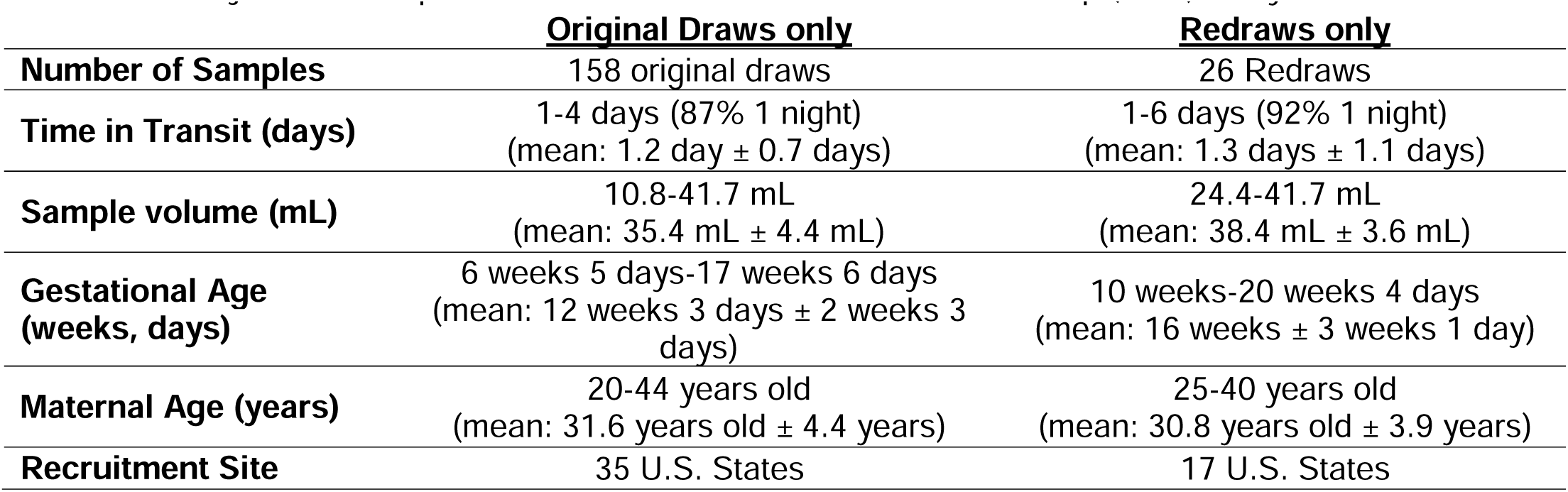
Subject and sample information associated with the no follow up (NFU) study cases.

|  | **<u>Original Draws only</u>** | **<u>Redraws only</u>** |
| --- | --- | --- |
| **Number of Samples** | 158 original draws | 26 Redraws |
| **Time in Transit (days)** | 1-4 days (87% 1 night)<br>(mean: 1.2 day ± 0.7 days) | 1-6 days (92% 1 night)<br>(mean: 1.3 days ± 1.1 days) |
| **Sample volume (mL)** | 10.8-41.7 mL<br>(mean: 35.4 mL ± 4.4 mL) | 24.4-41.7 mL<br>(mean: 38.4 mL ± 3.6 mL) |
| **Gestational Age (weeks, days)** | 6 weeks 5 days-17 weeks 6 days<br>(mean: 12 weeks 3 days ± 2 weeks 3 days) | 10 weeks-20 weeks 4 days<br>(mean: 16 weeks ± 3 weeks 1 day) |
| **Maternal Age (years)** | 20-44 years old<br>(mean: 31.6 years old ± 4.4 years) | 25-40 years old<br>(mean: 30.8 years old ± 3.9 years) |
| **Recruitment Site** | 35 U.S. States | 17 U.S. States |

The **FU series** was designed to determine the performance of the Luna test compared to CVS or amniocentesis results on the same cases. A total of 243 blood samples (7 unusable) were received from three recruitment sites: Columbia University Irving Medical Center (Dr. Ronald Wapner), Icahn School of Medicine at Mount Sinai (Dr. Joanne Stone), and Yale School of Medicine (Dr. Audrey Merriam). Samples were collected between Aug. 2021 and Sep. 2022 and an overview of sample characteristics is shown in Table 3, insert table. Details of cells recovered are provided in Table S1, insert table. Any woman planning to undergo CVS or amniocentesis at these sites was eligible for participation, and the indications for performing testing were heterogeneous. Many had increased risk including cytogenetic risk, monogenic risk, advanced maternal age, or an abnormal ultrasound. In all cases, blood was drawn prior to the invasive procedure. The results from CVS or amniocentesis served as accepted standard of care data for comparison to Luna test results. There was no attempt for rapid results, and no Luna results were returned to patients. This comparison would allow us to establish accuracy, specificity, sensitivity, positive predictive value (PPV), and negative predictive value (NPV) of the Luna test relative to CVS/amniocentesis (CVS/amnio).

**TABLE 3.** Subject and sample information associated with the 243 follow-up (FU) study cases.

|  |  |
| --- | --- |
| **Number of Samples** | 243 |
| **Time in Transit (days)** | 1-5 days (84% 1 night)<br>(mean: 1.3 days ± 0.8 days) |
| **Sample volume (mL)** | 11.6-41.2 mL<br>(mean: 33.2 mL ± 6.0 mL) |
| **Gestational Age (weeks, days)** | 10 weeks 1 day-25 weeks 1 days<br>(mean: 13 weeks 3 days ± 3 weeks 1 day) |
| **Maternal Age (years)** | 20-53 years old<br>(mean: 35.4 years old ± 4.6 years) |
| **Recruitment Site** | 67 Columbia, 133 Mt. Sinai, 43 Yale |

### 2.2 Protocol

Four 10 mL tubes of maternal blood (Cell-Free DNA BCT®, Streck) were collected carefully to maximize free flow with immediate inversion to avoid any clotting; blood was shipped overnight at ambient temperature on the day it was collected to Luna Genetics in Houston, TX. Blood was processed with a mild fixation of 2% paraformaldehyde for 10 min followed by a 5-fold volume increase resulting in a 0.08% Triton X-100 concentration for 4 min as a lysis procedure to eliminate red blood cells (RBC).^7,8^ Cells were permeabilized with a final concentration of 1X BD Perm/Wash Buffer 554723 (BD Biosciences) and stained with two antibodies to cytokeratin (Alexa Fluor 488 anti-cytokeratin [pan-reactive] C11 and BioLegend cat. No. 628608 and Alexa Fluor 488 anti-pan cytokeratin AE1/AE3 Life Technologies, cat. No. 53-9003-82) and CD45 (PE anti-human CD45 Antibody clone 2D1 BioLegend cat. No. 36851) as described previously.^7,8^ The stained nucleated cells were processed with a microfluidic sorting step to enrich ∼200 fold for CK-positive cells. Stained cells were loaded on to a Namocell Namo 3 or Hana instrument. In these systems, cells flow through a microfluidic channel and pass a laser at high density, and target cells are enriched through dispensing of a 1uL droplet upon detection by the laser. Cells with the highest CK staining intensity were collected and the enriched cell fraction was loaded into CyteSlides (RareCyte, Seattle, WA) with DAPI and subjected to automated cell scanning using a CyteFinder/CytePicker (RareCyte). Candidate cells were reviewed in a gallery (Figure S1), and presumptive fetal trophoblasts were manually picked into microfuge tubes using a micromanipulator within the CytePicker. Occasionally, cells were picked as trophoblast doublets, on the assumption that they were previously adjacent cells in the placenta. Individual cells or doublets were subjected to whole genome amplification using the PicoPLEX® Single Cell WGA Kit (Takara), and a DNA library suitable for NGS was prepared using the Nextera XT DNA Library Preparation Kit (Illumina). DNA was sequenced (single-end, read length of 100 bp) to obtain 1-5 million reads on an Illumina MiniSeq or NextSeq 2000, and the read data was analyzed using NxClinical (NxC) software (BioDiscovery). Up to five cells were analyzed for each case of singleton fetus (additional cells were analyzed in cases of twin pregnancy or suspected mosaicism, when possible). All single cell copy number profiles were plotted and analyzed against both a male and female reference data set. All cells were plotted and analyzed against a male and a female control with the male reference used in all figures. Reference sets controls were prepared from a pool of 10-20 male or female trophoblasts from low-risk pregnancies where no copy number abnormalities were evident. All cells were examined using both log ratio and copy number settings in NxC, with visual analysis performed on one chromosome at a time, noting any major deflections from normal copy number. For deletion/duplication analysis, the highest quality cell was selected, and each call made by the software was manually examined for assignment as a real copy number variant (CNV) versus technical artifact. Subsequently, each additional cell was reviewed in the same manner, checking for concordance against the highest quality cell and excluding calls that were deemed technical artifact. Preliminary categorical assignment of each CNV call was made by a genomics specialist (see 2.3 | Scoring Quality of NGS data). Copy number gains or losses meeting quality criteria were flagged for final review and interpretation by an ABMGG-certified lab director.

### 2.3 Fetal cell genotyping

Maternal genomic DNA was extracted concurrently from the blood sample and analyzed using a custom Illumina Global Screening Array v2 SNP array at a CLIA-accredited reference lab. The full SNP data for the mother was compared to limited SNP data for each cell from single cell NGS reads. Each cell was documented by genotyping to be of fetal and not maternal origin based on hundreds to thousands of SNP alleles present in the cell but not in the mother, whereas maternal cells had negligible such alleles. Egg donation and surrogate maternal variables were considered.

### 2.4 Scoring Quality of NGS data

Every cell subjected to NGS analysis was given one of three scores for deletion/duplication (del/dup) and/or aneuploidy as below by a genomics specialist:

1. Scorable for aneuploidy and 1.5 Mb del /2.0 Mb dup resolution
2. Scorable for aneuploidy only
3. Unscorable, not used for further analysis

Unscorable cells were either apoptotic, lost in processing, had very low mappable reads, or had data unsuitable for analysis for unknown reasons. Cells scorable for aneuploidy only resolution were often cells in S phase of the cell cycle where numerous small genomic segments not yet replicated cannot be distinguished from small deletions. Cells scorable for aneuploidy and 1.5 Mb del/2.0 Mb dup resolution had very high quality NGS data and very few putative gains or losses called by the NxC software. The final interpretation for the case was based on the cumulative findings in all the scorable cells. Each case was signed out as normal (no pathogenic copy number abnormalities detected) or as having a specific copy number abnormality. The numbers of cells with a score of 1 (scorable for aneuploidy and del/dup at 1.5/2.0Mb) or a score of 2 (aneuploidy only resolution) is specified in the figures and tables below. For ethics approval, the samples from the **Follow-Up series** were collected under a protocol approved by the Columbia University Medical Center IRB protocol AAAT6189 and WCG ID 20193442 and is ClinicalTrials.gov ID NCT04285814. The **No Follow-up series** was approved by the WCG IRB ID 20216940.

## 3 RESULTS

Blood was obtained from 158 low-risk pregnancies not undergoing an invasive procedure (**NFU series**), and from 243 pregnancies scheduled for CVS or amniocentesis (**FU series**). Additionally, single cells from seven chromosomally abnormal cell lines were processed through the same workflow to generate spike-in data. Figure 1A shows four normal trophoblast cells from one singleton pregnancy and Figure 1B shows single abnormal trophoblast cells, each from a different singleton pregnancy, demonstrating the robust detection of the three most common forms of autosomal trisomy (T13, T18, and T21). Two biological processes, apoptosis and S phase of the cell cycle, are found to interfere with cell analysis at a variable frequency (5-10% for each) in circulating trophoblasts. Figure S2A demonstrates a case with two normal cells and one apoptotic cell. Figure S2B shows cultured normal lymphoblasts (GM12878) in G1, S, and G2/M phases of the cell cycle analyzed with FACS to document DAPI intensity. Figure S2C demonstrates two cells from a case with trisomy 16; the trisomy is detected in both cells, although one of the cells is in S phase.

**FIGURE 1.**
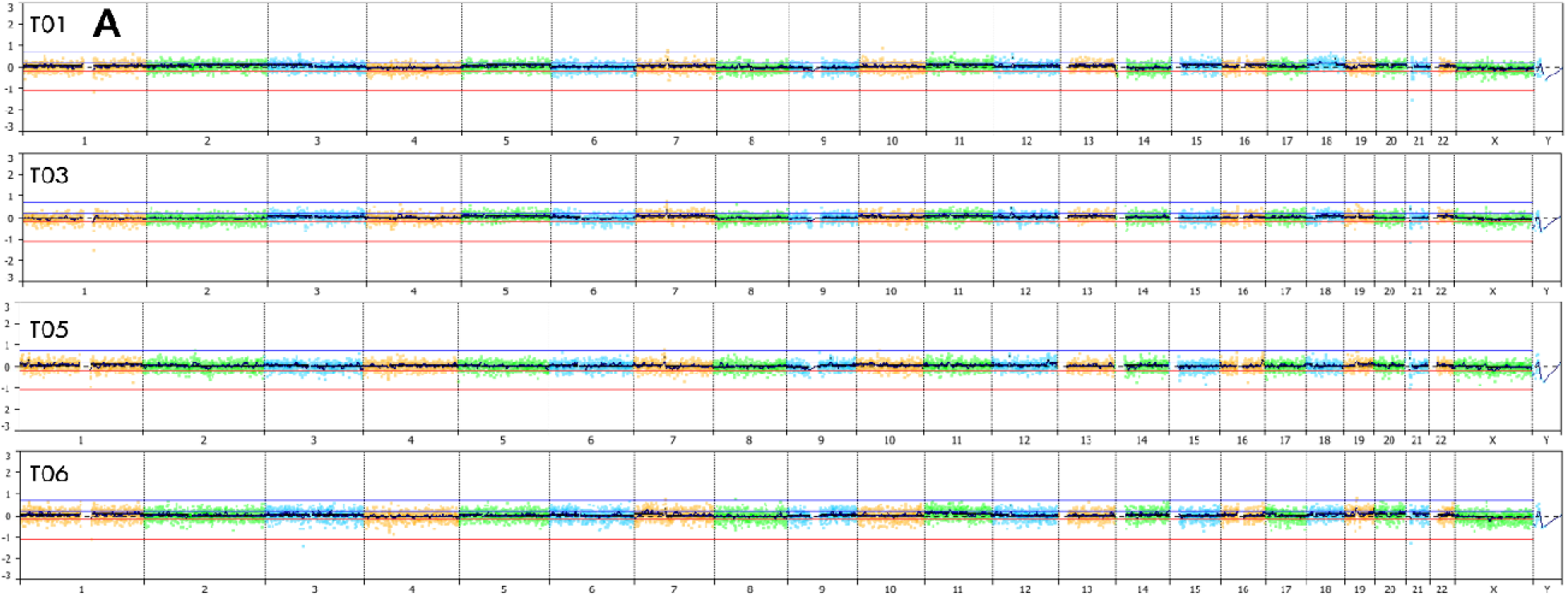
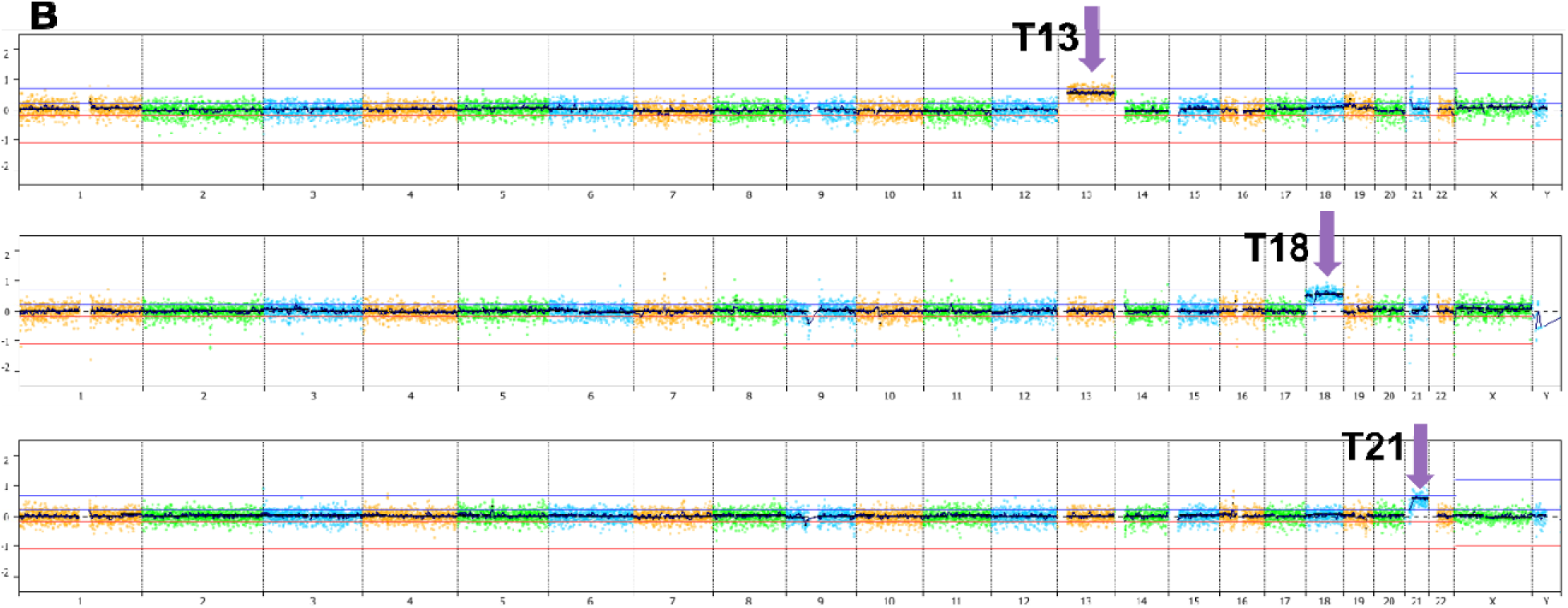
Examples of normal and abnormal cells analyzed with the single cell prenatal genetic test. Panel A includes four cells from a singleton pregnancy with a normal male fetus compared to a male control using a log ratio plot. A genomic copy number plot for all chromosomes is shown from chromosome 1 on the left to X and Y on the right using the NxClinical software. No gains or losses of copy number above reporting cutoff are observed in any cells. T01 to T06 identifies specific trophoblasts from this case. Panel B shows plots from three different cases for the three most common forms of trisomy as indicated. All are plotted against a male control; the T13 and T21 fetuses are male and the T18 female. Each plot shows the clear gain in copy number for the relevant chromosome. All displays of NxClinical data are in the Log Ratio format.

### 3.1 Analysis of 158 no-follow-up (NFU) samples from 35 states

For the **NFU series**, there were 158 first draw samples and 26 redraw samples from 158 pregnancies. The overall information for NGS scorable cells is shown in Figure 2A indicating the number and quality of trophoblasts for each case. Focusing on the combined data from first draw and redraws, one or more scorable cells were obtained for 91% of samples so that a report with results could be issued for those cases, and 9% of cases would have a report of “No Result-No scorable cells” (Table 3). Under ideal circumstances, the success might be slightly better because we did not request redraws in the first 40 cases, and not all women who were asked for a redraw gave a second blood sample. At least one cell yielded high-resolution data (score 1) in 75.9% of cases, and in 15.8% of cases the cells were scoreable for aneuploidy only (score 2) (Table 4, insert table). When a redraw was performed, at least one scorable cell was obtained in 77% of samples, and 30% of cases went from having only score 2 cells to having at least one score 1 cell. It is of interest to compare success in recovering scorable cells as related to gestational age as shown in Figure 2B. From this data, we suggest that the optimal time to perform the Luna test is 9-14 weeks of gestation, though it is also feasible to perform the test as early as 8 weeks or as late as 22 weeks or later. If cells are obtained, the Luna test is reliable, but there is a slightly increased risk of failure to obtain scorable cells outside of the 9-14-week range.^10,13^

**FIGURE 2.**
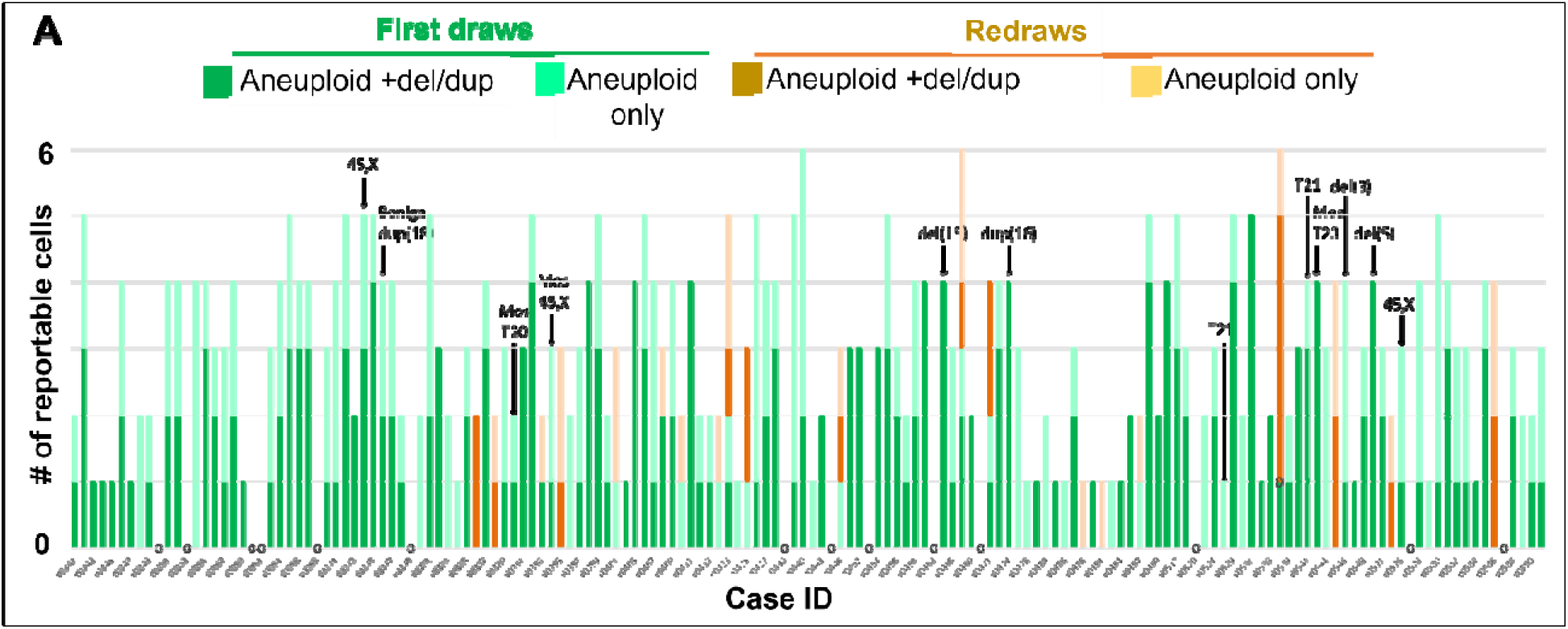
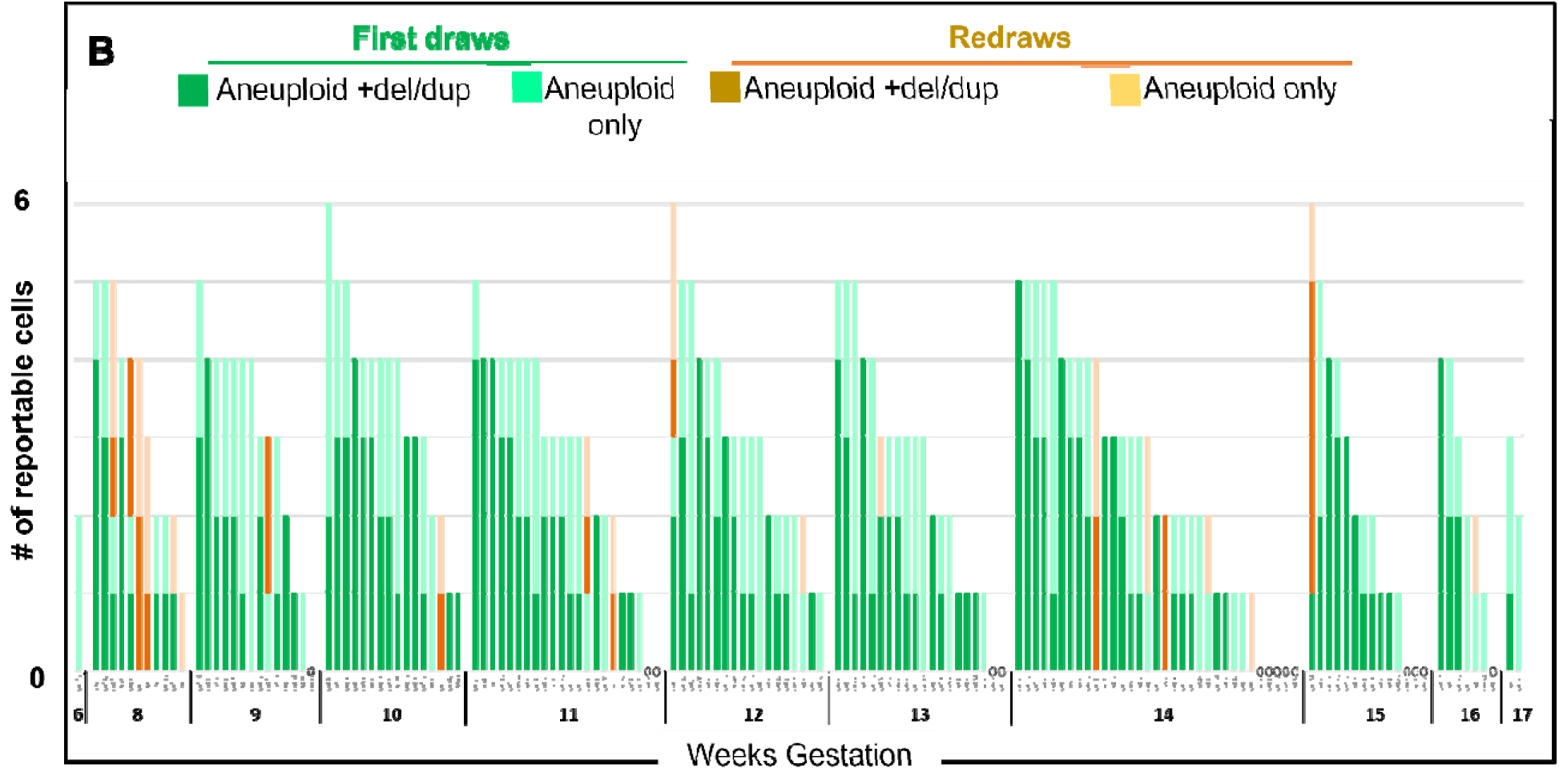
Cell analysis for 158 cases from across the US with no follow-up CVS or amniocentesis. In panel A, cases are plotted by date of blood collection between Jul. 2021 on the left through Jan. 2022 on the right. Each column is a case. Cells are designated by vertical bars as follows: Cells from first blood draw are in green and cells from redraw in brown. Cells scorable for aneuploidy and del/dup are in dark colors and cells scorable for aneuploidy only are in light colors. Cases with abnormalities are indicated in black font. In panel B, the same cases are plotted by gestational age at first blood draw.

**TABLE 4.**
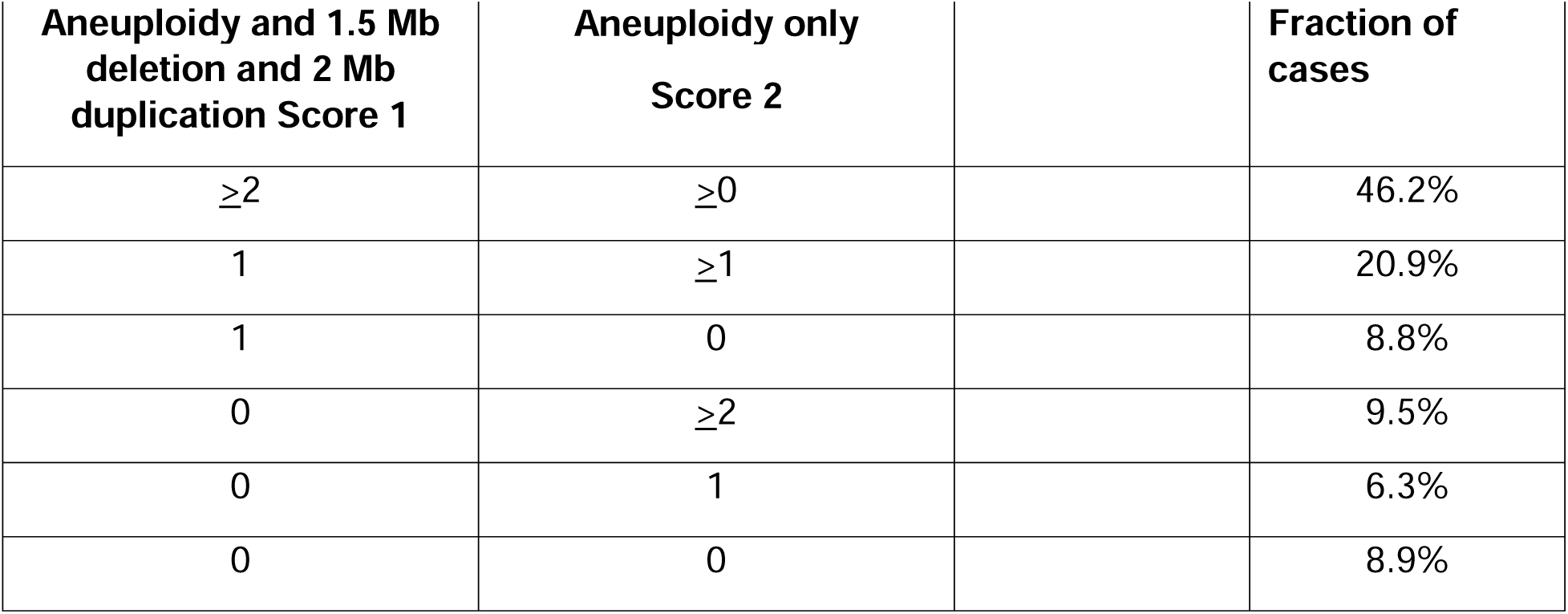
Reportable cells from 158 cases in the no-follow-up (NFU) series.

Aneuploidy abnormalities in the 158 **NFU series** included two with T21, three with 45,X (one mosaic), and two samples with mosaic trisomy 20 (Table 5), insert table. Subchromosomal findings (Table 5) included one each with deletion of 3p26.3pNot in 26.1, deletion of 15q11.2q13.1 (Prader-Willi/Angelman region), duplication of 16p13.11, and duplication of 18p11.32. Three of the subchromosomal events were interpreted as pathogenic or likely pathogenic, and one (dup18) was considered likely benign. Because of concordance of multiple cells within each abnormal sample (Figures. 3 and S3), we believe that these findings are real and not artifacts, but they could be subject to mosaicism and not present in CVS/amnio had they been performed. Follow-up contact was not available under IRB provisions, and thus results could not be tested further. The 3.9 Mb deletion 3p26.3p26.1 was interpreted as likely pathogenic.^18–20^ The 1.9 Mb duplication of 16p13.11 was interpreted as pathogenic with incomplete penetrance.^21–23^ The 6.3 Mb deletion of 15q11.2q13 was interpreted as pathogenic and consistent with a pathogenic class 1 deletion (BP1-BP3) causing either Prader-Willi or Angelman syndrome,^24,25^ which could not be distinguished according to parent of origin based on the information available. Despite the concordance of multiple cells within each abnormal sample (Figures. 3 and S3), follow-up contact was not available under IRB provisions and thus results could not be compared to CVS/amnio.

**FIGURE 3.**
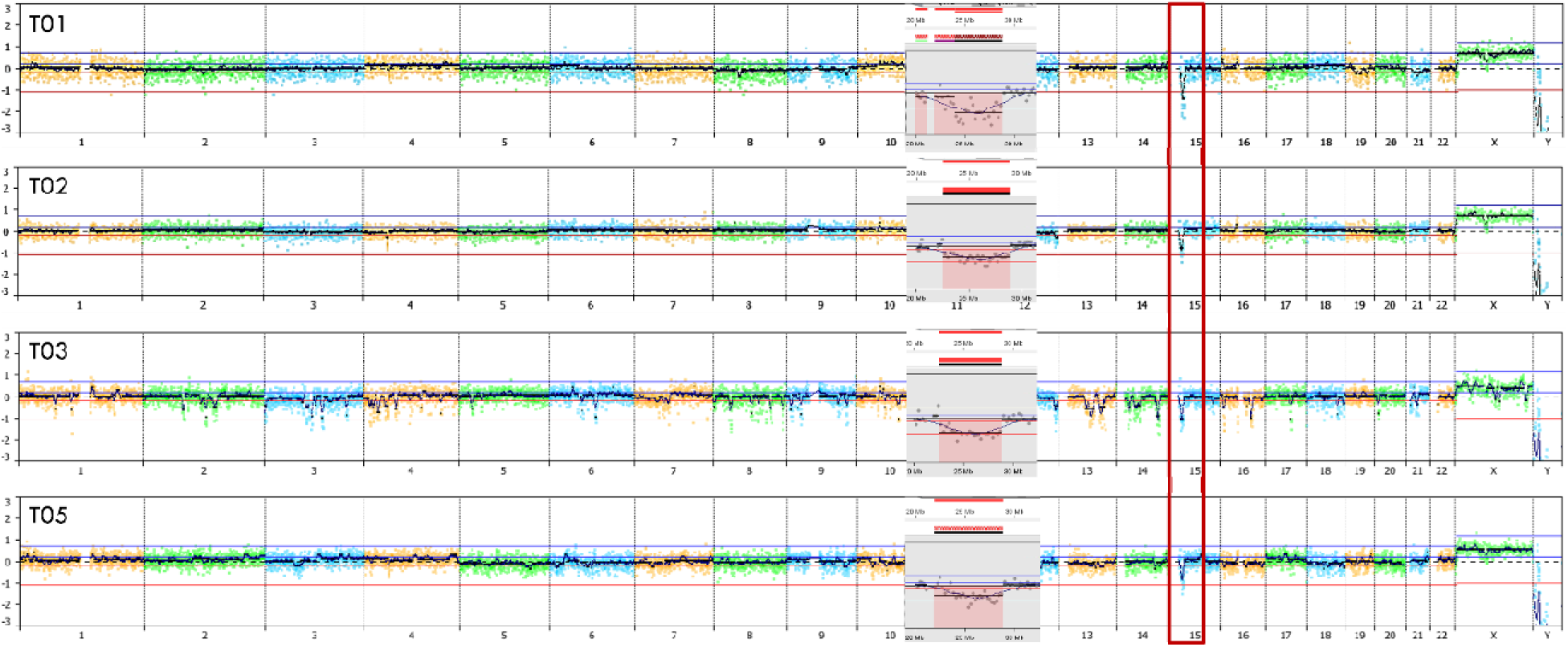
Detection of a Prader-Willi/Angelman deletion in all four available cells. The fetus is female, and data is plotted against a male control. The deleted region is in the red box on the genomic plots. In the insets, the relevant region on chromosome 15 is zoomed in for each cell.

**FIGURE 4.**
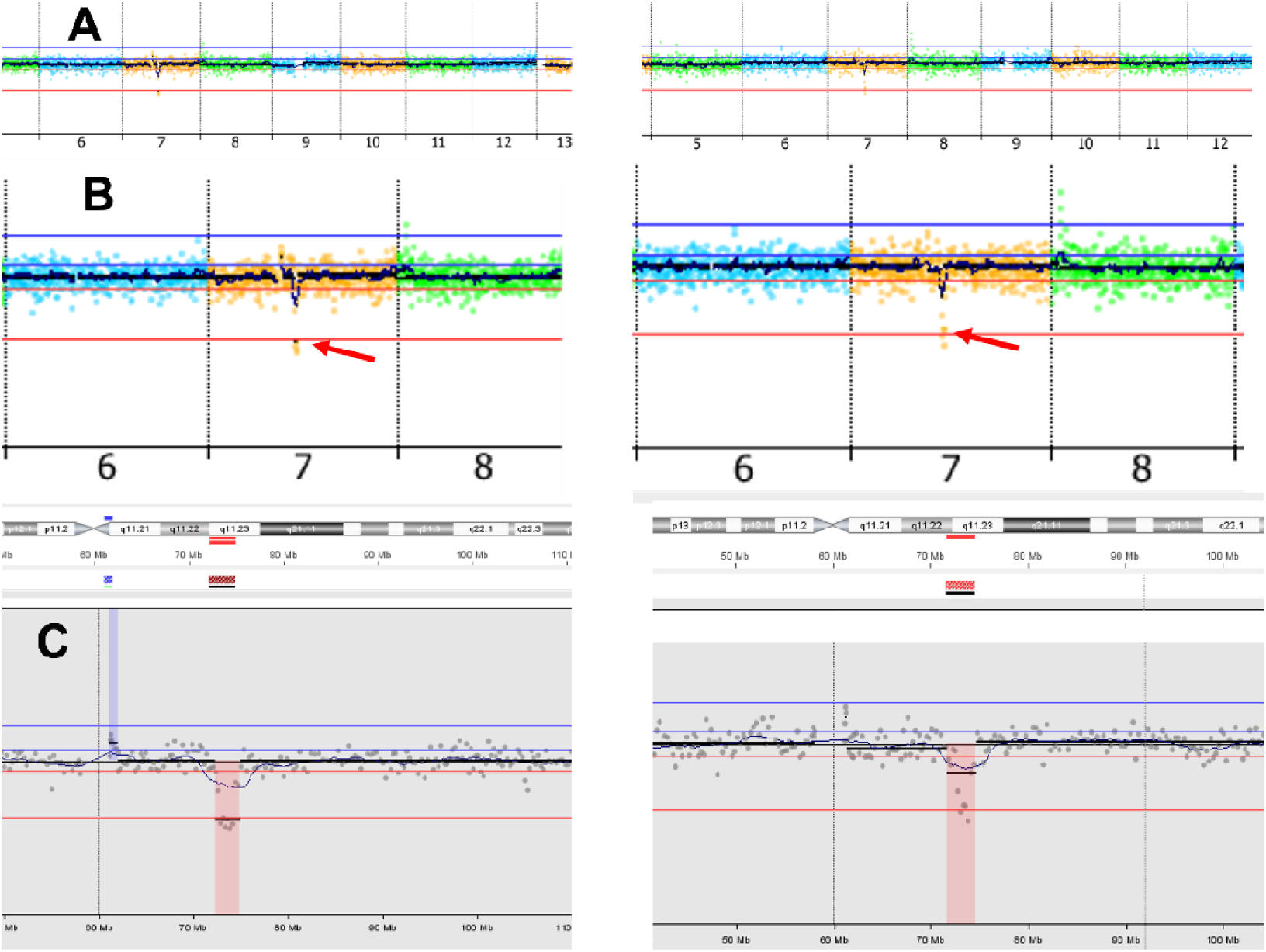
Detection of a Williams syndrome deletion (7q11.23) in two cells from a monochorionic twin pregnancy. Panel A shows a view of chromosomes 6 to 13 from genomic plots, panel B chromosomes 6 to 8, and panel C a subregion of chromosome 7.

**TABLE 5.** Chromosomal abnormalities† in 158 pregnancies in the no follow-up (NFU) series.

| **Result** |  | **# of cases** | **% of cases** |
| --- | --- | --- | --- |
| **Normal** | 46,XX or 46XY | 134 | 84.8 |
| **Abnormal** | 45,X (1 mosaic) | 3 | 1.9 |
|  | Trisomy 21 | 2 | 1.3 |
|  | del(3)(p26.3-p26.1) (3.9Mb) | 1 | 0.6 |
|  | del(15)(q11.2 - q13.1) (6.3Mb) | 1 | 0.6 |
|  | dup(16)(p13.11) (1.9Mb) | 1 | 0.6 |
|  | Trisomy 20 (both mosaic) | 2 | 1.3 |
| **No scorable cells** | No results | 14 | 8.9 |
| **Total # of cases** |  | 158 |  |

†Dup 18p11.32 nonpathogenic not included.

### 3.2 Concordance between cells in a case

Of the 158 singleton **NFU** pregnancy cases, 118 cases (75%) had two or more reportable cells. This count excludes three mosaic cases (two T20 and one 45,X) which by definition have cells of differing results. All cells (total 404 cells) within each of those 118 cases showed concordant results for aneuploidy. There were 74 cases with two or more cells of high quality (score 1). All 210 of these score 1 cells agreed as to the presence or absence of any pathogenic changes. When scoring for clinically significant copy number abnormalities, the concordance between cells within the same sample was 100% (excluding the three samples with placental mosaicism).

### 3.3 Analysis of 243 follow-up (FU) samples with CVS and amniocentesis comparison

The results for the 243 cases in the **FU series** are summarized in Table 6A, insert table. For 160 singletons pregnancies (83 female and 77 male), Luna results were normal, and sex agreed with CVS/amnio results. For 14 aneuploidy cases, nine T21, three T18, one T13, and one XXX, Luna test results agreed with those from CVS/amnio . For one case, the chromosome analysis of CVS tissue showed T21 in all 20 cells analyzed with four cells also showing T8 while T8 was not detected in any cells analyzed by the Luna test. In one case of monochorionic twins, results indicated a Williams syndrome deletion in all studies, but neither the Luna test nor CVS could distinguish one twin from the other. Amniocentesis was not performed, and post-termination studies were not performed. Based on the monochorionic placenta including confirmatory vascular studies, both twins likely carried the deletion, but separate studies of each twin were not performed. One case in the “incomplete information” group showed a deletion of 1.5 Mb at 17p12 consistent with a diagnosis of hereditary neuropathy with liability to pressure palsies (OMIM 162500) in five cells on the Luna test. Two cells were scored as aneuploidy only due to quality thresholds, but the deletion was still visually evident. Unfortunately, this finding could not be confirmed because only karyotype analysis was performed on the cultured CVS, which would not have detected a deletion of this size. No scorable trophoblasts were recovered in 39 of 236 usable samples.

**TABLE 6A.**
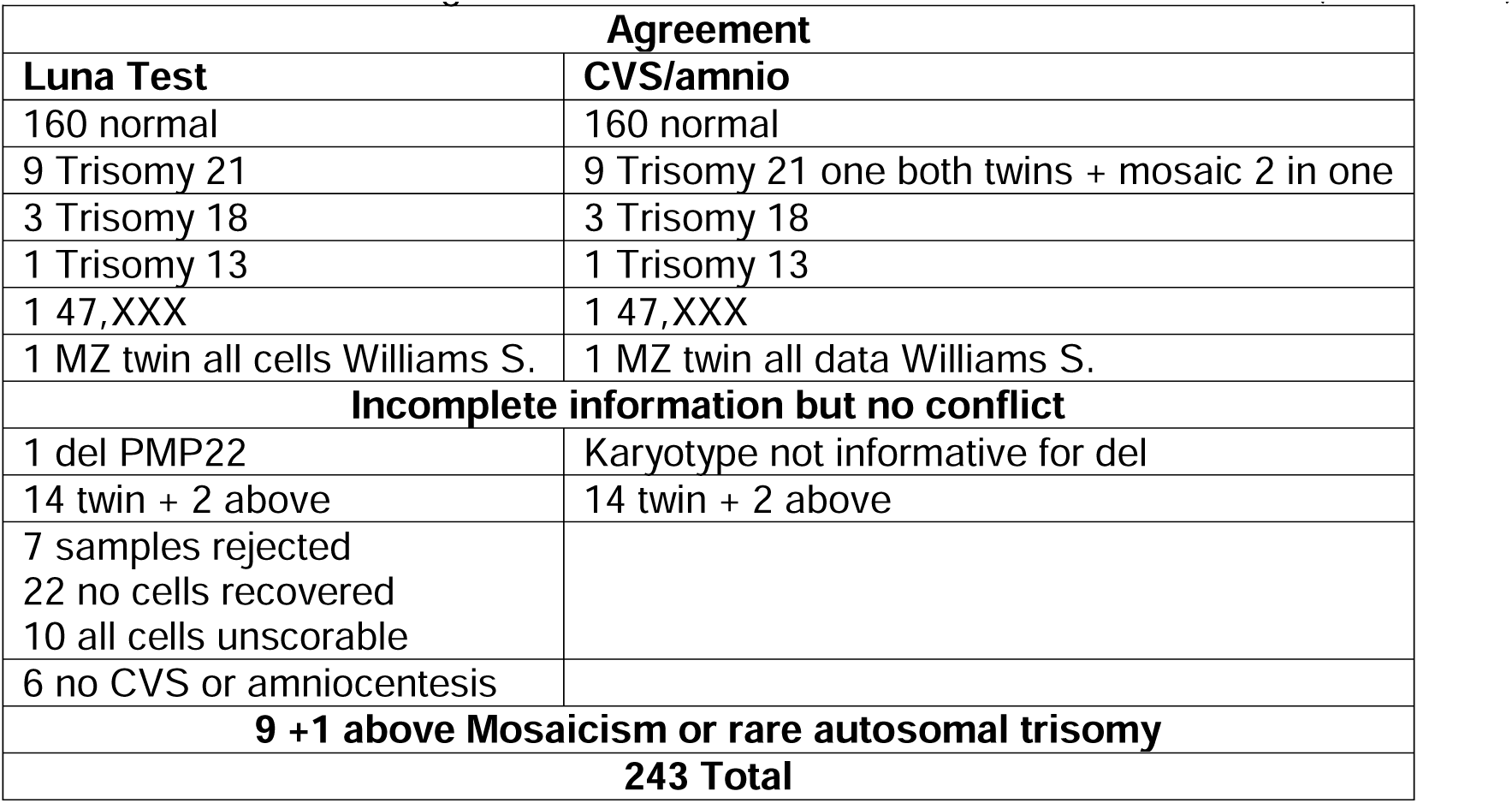
Cases with agreement of Luna test with CVS or amniocentesis (187 cases)

There were 10 cases with differences between the Luna test and CVS/amnio data (Table 6B, insert table), all of which were compatible with placental mosaicism. We interpret all results from all procedures to be real and accurate with findings compatible with placental mosaicism. We would not designate such cases as false positive or false negative, but rather conclude that the correct diagnosis is placental mosaicism with differing but correct results in two samples. Placental mosaicism occurred in 7 of 236 Luna cases (3.0%) and in 3 of 188 (1.6%) CVS cases (total 4.6%). In one noteworthy case, the Luna test detected T13 in four out of 4 trophoblast cells, but a karyotype on cultured cells from amniocentesis was normal (Table 6B). This case documents why the Luna test is not diagnostic for aneuploidy (see Discussion). Interestingly, no case showed placental mosaicism in both the Luna test and CVS, suggesting that neither the current Luna test alone nor CVS alone detect all placental mosaicism for aneuploidy.

**TABLE 6B.**
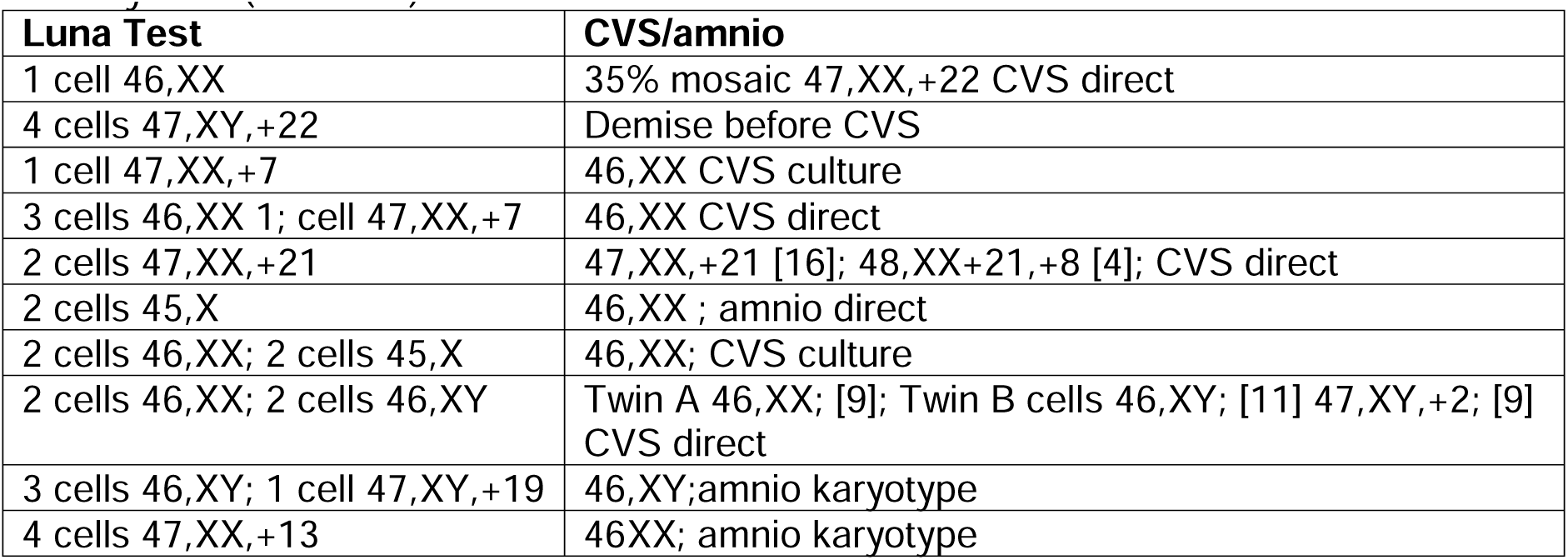
Cases with differences compatible with placental mosaicism or rare autosomal trisomy/RAT (10 cases)

There were six cases (Table 6C, insert table) with a Luna test result but no CVS/amnio result. These included three trisomy cases (one T13, one T21, and one T22) in which there was a fetal demise before a scheduled CVS/amnio. There were two Luna cases (one T18 and one normal Luna result but abnormal ultrasound heart defect) where pregnancies were terminated based on existing pathological information without CVS/amnio. There was one normal Luna case with a failed CVS.

**TABLE 6C.** Cases with Luna result but no CVS or amniocentesis result (6 cases)

| **Luna Test** | **CVS/amnio** |
| --- | --- |
| 4 cells 47,XY,+22 | Demise before amnio |
| 1 cell 47,XX,+18 | cfNIPT T18 declined CVS termination |
| 5 cells 46,XX | Failed CVS |
| 4 cells 47,XY,+13 | cfNIPT T13 demise before CVS |
| 1 cell 47,XY,+21 | cfNIPT T21 demise before CVS |
| 2 cells 46,XX | Termination based on abnormal US |

Details for 15 twin cases are provided in Table S2, insert table. Based on CVS/amnio data, there were six cases of opposite sex twins, and the Luna test found cells of both sexes in four cases but identified only male cells in one case and only female cells in another case. There were four cases of female/female twins, and the Luna test could not distinguish one twin from the other including one case of Williams syndrome deletion likely in both twins. There were two cases of male/male twins, and the Luna test could not distinguish one twin from the other including one case with CVS/amnio showing mosaic T2 in one twin but no T2 in the other twin and no T2 in the Luna cells. There were no unexpected sex differences, such as male cells identified by Luna in confirmed female/female twin pairs or vice versa.

### 3.4 Conclusions for CVS/amnio comparison

The Luna lab was blinded to all results of CVS/amnio until it had submitted reports to a joint database. In general, the agreement between the Luna test and CVS/amnio for singleton pregnancies was 100%, with the caveat of 10 cases involving placental mosaicism that led to differing results. In addition, there were two cases reported as normal by Luna which were reported as having a balanced translocation by CVS/amnio, and these were counted as agreement. There was one case where Luna detected a 17p12 deletion, but only karyotype was performed on the CVS, and this was not counted as a disagreement. There were 39 cases where no cells were recovered (Table 6A) by the Luna test, but data was available for CVS/amnio. Of the 39, seven did not meet sample acceptance criteria and were rejected, 22 failed to recover cells, and in 10 cases cells were recovered and subjected to NGS, but all cells were unscorable. For the 22 with failure to recover cells, we reviewed whether these were associated with any pathological state. There were 18 normal results, two T21, one mosaic 45,X, and one unbalanced translocation. This was not significantly different from the spectrum of cases where cells were recovered. Thus, a failure to recover cells with the Luna test was not significantly associated with abnormal cytogenetic abnormality by CVS/amnio. For the 204 cases where Luna results were informative, 81 (40%) of the Luna samples could be scored for aneuploidy only and not for deletion/duplication. No deletion/duplication findings by CVS/amnio were missed by the Luna test.

### 3.5 Calculation of sensitivity and specificity for Luna test versus CVS/amnio

For the 243 samples received (236 usable), 160 agreed for normal, 14 agreed for trisomy, and 1 agreed for twins with Williams syndrome. Although there were no disagreements for the twin cases, same sex twins were not distinguished from each other by the Luna test. Thus, only twins where both sexes were identified in the Luna test and in the follow-up test were counted for these sensitivity and specificity calculations. If cases interpreted as placental mosaicism were excluded (Fig. 5A), the accuracy, sensitivity, specificity, PPV, and NPV were all 100%, but the total number of cases is limited, and the confidence intervals are significant. If one counts mosaic cases as false positives or false negative, the values for accuracy, sensitivity, specificity, PPV, and NPV are all less than 100% as shown in Fig. 5B. Although one Luna result had all four cells with T13 and a normal amnio result might be called a false positive, we would describe this as a case with 4/4 cells showing T13 in the placenta and no abnormal cells on amniocentesis. The consequences of the T13 in the placenta may have implications for the course and outcome of the pregnancy. All other Luna positives that were interpreted as placental mosaicism involved 45,X or rare autosomal trisomy (RAT) where the possibility of mosaicism is high and would likely be followed up by CVS/amnio. For the three cases where mosaicism was absent on the Luna test but present on direct CVS, these emphasize that the fewer the cells recovered by the Luna test, the higher the probability of missing placental mosaicism. Of course, the Luna test does not detect type 2 mesenchymal mosaicism.

**FIGURE 5.**
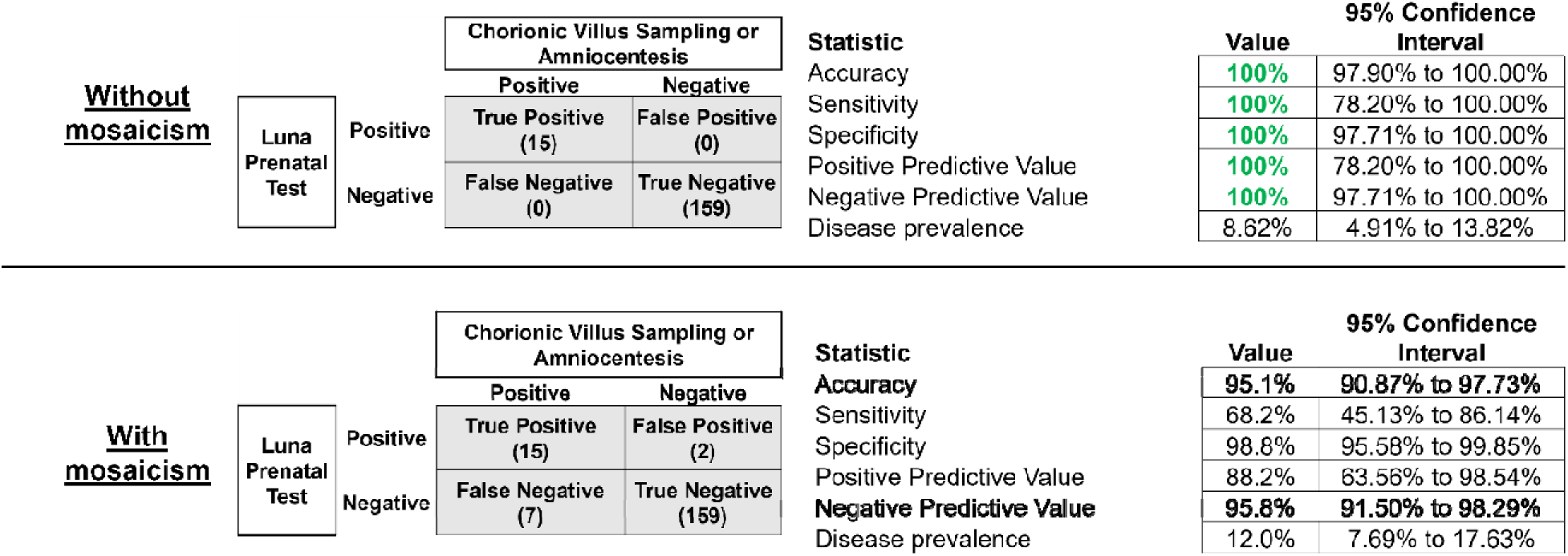
Statistical analysis of comparison of Luna test to CVS and amniocentesis.

### 3.6 Clinical and Analytical Performance with spike-in cells

Give that it is not feasible to obtain samples from women carrying fetuses with a wide range of deletions and duplications, we used human cultured cells (lymphoblast or fibroblast obtained from biobanks at the Coriell Institute for Medical Research) with known aneuploidy or deletion or duplication as shown in Table 7, insert table. Out of a total of 148 scorable cells, known abnormalities were detected and called by the NxC software in every cell except one unscorable cell. An example of the data for a 1.5 Mb deletion cell line is shown in Figure S4. Thus, the analytic sensitivity and specificity were near 100% with the expected finding being reported in every cell. This included inter-prep and intra-prep replicates.

**TABLE 7.** Cell line IDs and associated abnormalities.

| **<u>Coriell ID</u>** | **<u>Cell Line ID</u>** | **<u>Abnormality</u>** | **<u>Size</u>** |
| --- | --- | --- | --- |
| GM03102 | C0001 | 47,XXY | whole chromosome |
| GM03538 | C0002 | Trisomy 18 | whole chromosome |
| GM03716 | C0003 | Trisomy 21 | whole chromosome |
| GM22049 | C0004 | 47,XY,+idic(15)(q13) | 12 Mb |
| GM23007 | C0005 | del(1)(p36.33-p36.22) | 9.2 Mb |
| GM17942 | C0006 | del(22)(q11.21) | 2.8 Mb |
| GM24084 | C0007 | del(17)(p12) | 1.5 Mb |

The clinical and analytical performance characteristics of the Luna test based on results with spike-in cells were evaluated for accuracy, precision, sensitivity, and specificity. Overall, clinical sensitivity, specificity, PPV, and NPV were each 100% for all the observed conditions. Additionally, reproducibility and repeatability studies were performed using the same sample set to demonstrate precision of results across multiple replicates of the assay (inter-run precision). Three replicates of a single sample on a single run were performed to demonstrate precision of test results under the same operating conditions (intra-run precision). The observed results from each of the runs performed were 100% concordant. In addition, we performed a Limit of Detection Analysis by spiking in Coriell cell lines harboring characterized copy number abnormalities, which confirmed that our clinical Luna test can detect copy number losses down to a deletion of 1.5 Mb or a duplication of 2.0 Mb (Table 7 and Figure S4).

## 4 DISCUSSION

The Luna single cell noninvasive prenatal genetic test (the Luna test) has significant advantages compared to cfNIPT. The Luna test can determine a high-resolution copy number genotype on one or more circulating trophoblasts from a pregnancy. In this regard the data is very similar to those long familiar from chromosomal microarray analysis. The validation data reported here indicates that the test can accurately detect deletions and duplications at a resolution of 1.5 Mb and 2.0 Mb, respectively, which is superior to any deletion/duplication detection method using cfNIPT. Many sources of false positive results with cfNIPT relate to an abnormal maternal genotype, and these effects are avoided by the Luna test. False positive results with cfNIPT lead to unnecessary anxiety and unnecessary CVS and amniocentesis, and sometimes unnecessary termination of pregnancy. Another source of variation with cfNIPT is low fetal fraction or difficulty in setting cutoff values to perfectly separate normal and abnormal results. In the Luna test, each cell is assigned a normal or abnormal copy number genotype or is designated unscorable. The Luna test can be performed as early as 8 weeks gestation and is maximally successful by 9 weeks. For cfNIPT, fetal fraction decreases with increasing maternal weight,^26^ while maternal weight appears to be minimal or absent as a modifying factor for single cell testing;^27^ this is consistent with our experience (data not shown). For cfNIPT, maternal autoimmune disease and therapy with low molecular weight heparin decrease fetal fraction,^26^ and we speculate that these variables would not affect single cell testing. Although not shown in this report, preliminary data supports the feasibility of using SNP genotyping to distinguish cells from different fetuses in same sex dizygotic twins. In comparing the Luna test to cfNIPT, CVS, and amniocentesis, the Luna test can be performed substantially earlier than amniocentesis and slightly earlier than cfNIPT and CVS. Notably, it can be performed in the window from 13-14 weeks where neither CVS nor amniocentesis are available.

The weaknesses of the Luna test include failure to recover scorable trophoblasts in some cases, and recovery of cells that can be scored for aneuploidy only but not for del/dup. The failure to recover cells of sufficient quality can be addressed in part through a repeat blood draw at 14-15 days after the initial phlebotomy. In 26 cases with 0-1 cells recovered, a redraw was processed and resulted in additional scorable cells in 77% of cases. There is no evidence that failure to recover cells is an indication of any abnormality with the pregnancy and may occur more frequently in normal pregnancies, as the number fetal cells is reported to be increased with some aneuploid fetuses.^28^ Even when an optimal number of scorable cells is recovered, they may be few in number such as three or less, which may cause failure to detect aneuploidy mosaicism, especially for RAT, 45,X, andT13 Failure of the Luna test to detect rare autosomal trisomy was seen in two cases in Table 6B where only one cell was recovered with the Luna test. At present, a relatively large blood volume (40 mL) is being collected, but this remains a superior sample collection method when compared to the risks of CVS or amniocentesis. The inability to detect the smallest deletions and duplications such as single exon events remains a weakness relative to CVS and amniocentesis, but this could be addressed by deeper sequencing of single cells. The current cost of the test limits its uptake in many healthcare settings.

The **FU series** demonstrates complete agreement when the Luna test and CVS or amniocentesis are performed on the same singleton pregnancy, apart from 10 cases of placental mosaicism. The Luna test is not diagnostic for aneuploidy or deletions and duplications as is definitively shown by one case with four T13 cells by the Luna test and a normal karyotype by amniocentesis. This is consistent with recommendations from professional organizations that a final diagnosis should not rely on testing of trophoblasts alone (Association for Clinical Cytogenetics [ACC] Prenatal Diagnosis Best Practice Guidelines 2009). Therefore, it is recommended to confirm a positive NIPT result with invasive prenatal testing, preferably by amniocentesis (a joint European Society of Human Genetics [ESHG]/American Society of Human Genetics [ASHG] position statement).^29^ A publication by Van Opstal and Srebniak^30^ provides excellent guidance on the relative value of CVS (especially mesenchymal core analysis) and amniocentesis to confirm positive aneuploidy results based on cfNIPT. Because cfNIPT and the Luna test depend entirely on trophoblast DNA, these authors argue that “This is the reason that cfNIPT will and can never be a diagnostic test.” The circumstance where follow-up testing is most compelling is when data suggests an abnormality, and a termination or other therapeutic intervention is being considered. Compelling ultrasound data or follow-up CVS or amniocentesis confirmation are essential before proceeding. Placental mosaicism for deletions and duplications does occur, but its frequency is uncertain,^31–33^ and apparent pathogenic deletions and duplications should be confirmed by CVS or amniocentesis if any pregnancy management decisions are to be made. Mosaicism may occur for supernumerary, marker, or acentric chromosomes^31^ and possibly for any copy number variation. Performing a CVS or amniocentesis following a normal Luna test is not likely to be performed unless there is some other evidence of risk (e.g., abnormal ultrasound or inherited monogenic risk), although there is a measurable risk of missing placental mosaicism depending on the number of trophoblasts studied. If only one or two cells are reported or if no cell is scorable for del/dup, CVS or amniocentesis remain an option.

Many possible improvements can be envisioned for cell-based noninvasive prenatal testing. The potential to improve the recovery of trophoblasts and/or recovery of other fetal cell types from a given volume of blood is suggested by credible reports of 3-6 fetal cells per mL of maternal blood,^34^ compared to the recovery of 0.1-0.2 scorable cells per mL of maternal blood reported here and compared to 0.18/ cells per mL reported by the Menarini lab^13^ and 0.42 cells per mL (not corrected for scorability) reported by the ARCEDI lab.^8^ We observed a 9% failure rate of in the **NFU series** and 14% in the **FU series**, while Menarini^13^ reports a failure rate of 27-34%, and ARCEDI^11,14^ a failure rate of 7.6-10%. Hou et al., in a correction to a publication, reported 1.1-3.1 trophoblasts per mL of blood in six normal male pregnancies.^9,35^ Recovery of other cell types such as fnRBC, fetal lymphocytes, or fetal hematopoietic stem progenitor cells (HSPC) is theoretically possible and desirable, but the authors are not aware of reliable recovery of such cells in clinically usable numbers in the first trimester. Non-trophoblast cells would be especially helpful in addressing placental mosaicism. Processes for downstream analyses of recovered cells could also be improved. Particularly, cells in S phase are currently not scorable for small deletions. Expanded analyses of SNPs with attention to B-allele ratio and other forms of deeper NGS are likely to improve this limitation. We have observed that deeper sequencing of cells in S phase can provide improved del/dup detection (data not shown). Also, if the focus is on detecting inherited or de novo monogenic mutations, S phase cells are not at a disadvantage.

The use of cell-based noninvasive prenatal testing is impacted by placental aneuploidy mosaicism, but only a small fraction of pregnancies is involved (5.3% in this study). Placental mosaicism is classified as type 1 if confined to trophoblasts, type 2 if confined to the chorionic villus stroma, and type 3 if both trophoblast cells and the villus stroma are involved. Placental mosaicism is described as confined placental mosaicism (CPM) if limited to the placenta and absent in the embryo, or as true fetal mosaicism (TFM) if both embryo and placenta have abnormal cells. Since the Luna test utilizes trophoblasts for analysis, it will permit detection of types 1 and 3 mosaicism, but not type 2. It is likely that cfNIPT also detects types 1 and 3 but not type 2, since fetal DNA is derived from trophoblasts. CVS can detect all types of placental mosaicism with evidence that direct cell analysis queries primarily cytotrophoblasts, but long-term culture queries primarily mesenchymal cells. The data reported here suggests that neither the Luna test nor CVS detect all trophoblast mosaicism. We observed multiple examples of both Luna test-negative and CVS/amnio-positive and Luna test-positive and CVS/amnio-negative results. This is not surprising as placental mosaicism represents a complex sampling problem, and few cells are available in the Luna test. It is known that aneuploidy mosaicism is quite common in day 5 embryos generated in vitro.^36^ Based on this study, the occurrence of type 1 or 3 mosaicism in the placenta is not always detected by either the Luna test or CVS and is likely slightly higher than is often reported;^37^ here we observed 10 cases out of 188 potentially informative cases (5.3%).

There is some debate over whether it is desirable or undesirable to detect placental mosaicism for rare autosomal trisomy when present. Some laboratories study only cultured cells on CVS knowing that type 1 and/or /3 mosaicism will be missed. More recent publications emphasize the increased probability of various pregnancy complications involving mosaicism for RAT. The data reported here suggests that the frequency of detection of RAT may be higher than what is seen for types 1/3 on CVS. In such cases, we propose that the correct diagnosis is placental mosaicism, and all the observed data is true and accurate. More recent studies argue that detecting placental mosaicism for RAT is in the best interest for optimal management of a pregnancy.^38,39^ Such abnormalities carry risks of miscarriage, birth defects, and low birth weight. It has been suggested that such pregnancies should be followed more closely, for example by more frequent ultrasounds. However, the limited number of trophoblast cells recovered by the Luna test means mosaicism won’t always be detected when present.

Cell-based NIPT is an extremely promising approach to genetic prenatal diagnosis, although the current inability to recover 5-10 cells in >95% of cases limit its widespread adoption. If sufficient cells could be recovered, it is feasible to perform high-coverage whole genome sequencing (WGS) on trophoblasts^12^ raising the possibility of noninvasive trio WGS in the first trimester.

## ACKNOWLEDGEMENTS

We thank Faith Jacobs for performing DNA sequencing. Philip Reilly has played a leading role in many activities at Luna Genetics. We thank Patrick Koty and Noah Spies for critical reading of the manuscript.

## ETHICS STATEMENT

For ethics approval, the samples from the **Follow-Up series** were collected under a protocol approved by the Columbia University Medical Center IRB protocol AAAT6189 and WCG ID 20193442. The **No Follow-up series** was approved by the WCG IRB ID 20216940. (also in text)

## DATA AVAILABILITY STATEMENT

Raw data is not shared. Much of the processed data is available in figures and tables, especially Table S1. Additional processed data that support the findings of this study are available from the corresponding author [ALB} upon request.

---

Table S1 Excel Separate file

**Table S2.** Cases of twin pregnancies (15 cases)

| **Luna test number of cells** | **CVS/amnio** |
| --- | --- |
| Female 6 | Female/Female |
| Female 2 | Female/Female |
| Female 4 | Female/Female |
| Female 5 (all 7q11.2 del) | Female/Female (all 7q11.2 del) |
| Female 3/Male 1 | Female/Male |
| Male 5 | Male/Male (MCC* in A) |
| Female 4 | Female/Male |
| Female 1/Male 3 | Female/Male |
| Male 1 | Female/Male |
| Male 3 | Male/Male |
| Male 2 | Male/Male |
| Male 5 | Male/Male |
| Female 1/Male 3 | Female/Male |
| Male 2 | Male mono-di |
| Female 2/Male 2 (All normal) | Female/Male (Male mosaic +Chr:2) |

\*MCC, maternal cell contamination

**FIGURE S1 Example of trophoblast and maternal white blood cell staining.** One CK^+^/CD45-trophoblast (Arrow T) and two CK^-^/CD45+ maternal white blood cells (arrows WBC) Panel A CD45 staining and panel B cytokeratin staining.

**FIGURE S2.**
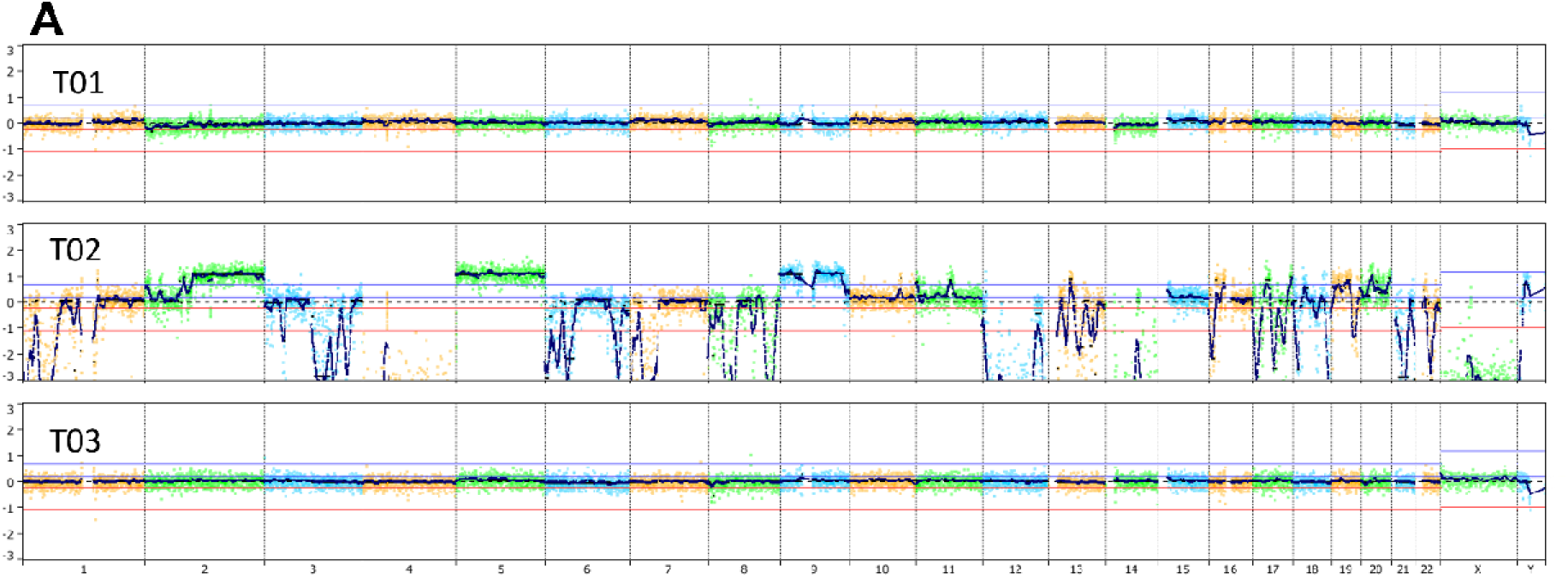
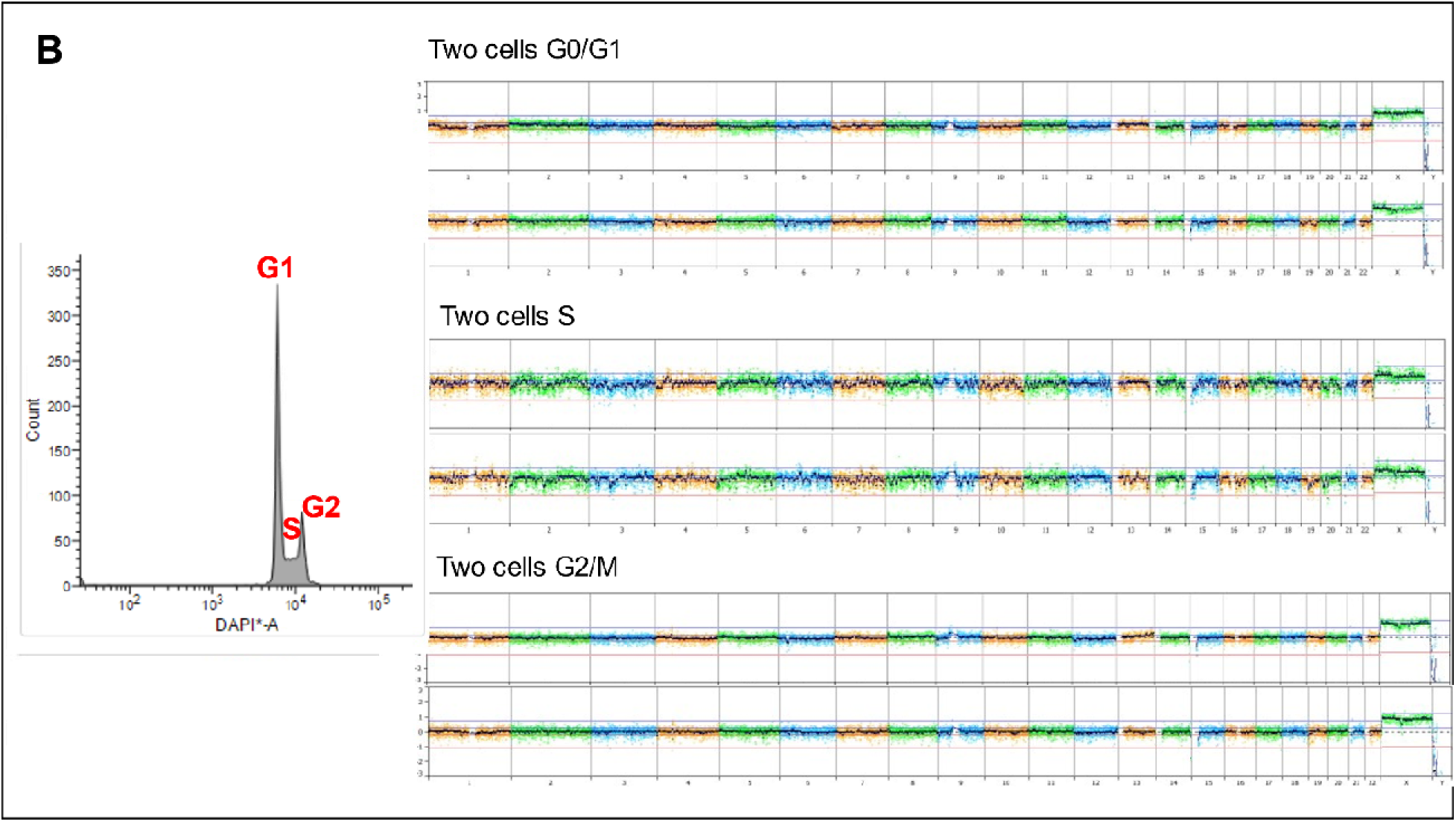
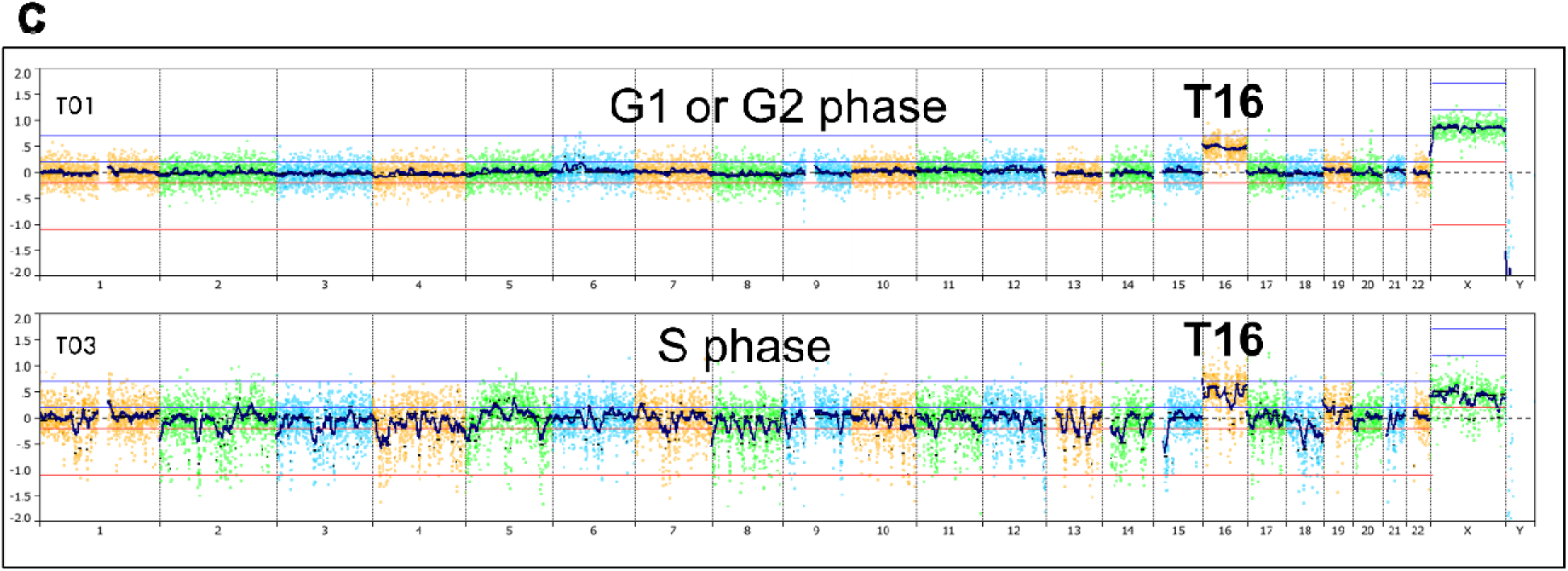
Examples of an apoptotic cell and cells in S phase. (A) Three cells from a normal male fetus. Cell T02 is apoptotic while T01 and T03 are not. (B) On the left is a plot of FACS analysis for DNA content (DAPI stain) of a normal female lymphoblast cell line plotted with male control. On the right is single cell analysis for two cells from each of the regions marked as G1, S and G2 in the left panel. In the S phase cells, many deletion-like events found across the entire genome represent late-replications regions. (C) Two cells are shown from a female fetus with trisomy 16 using a male control. Cell T01 is of better quality, but cell T03 is in S phase and shows widely variable copy number across the genome, reflecting early and late replicating regions, but the T16 is detectable in both cells.

**FIGURE S3.**
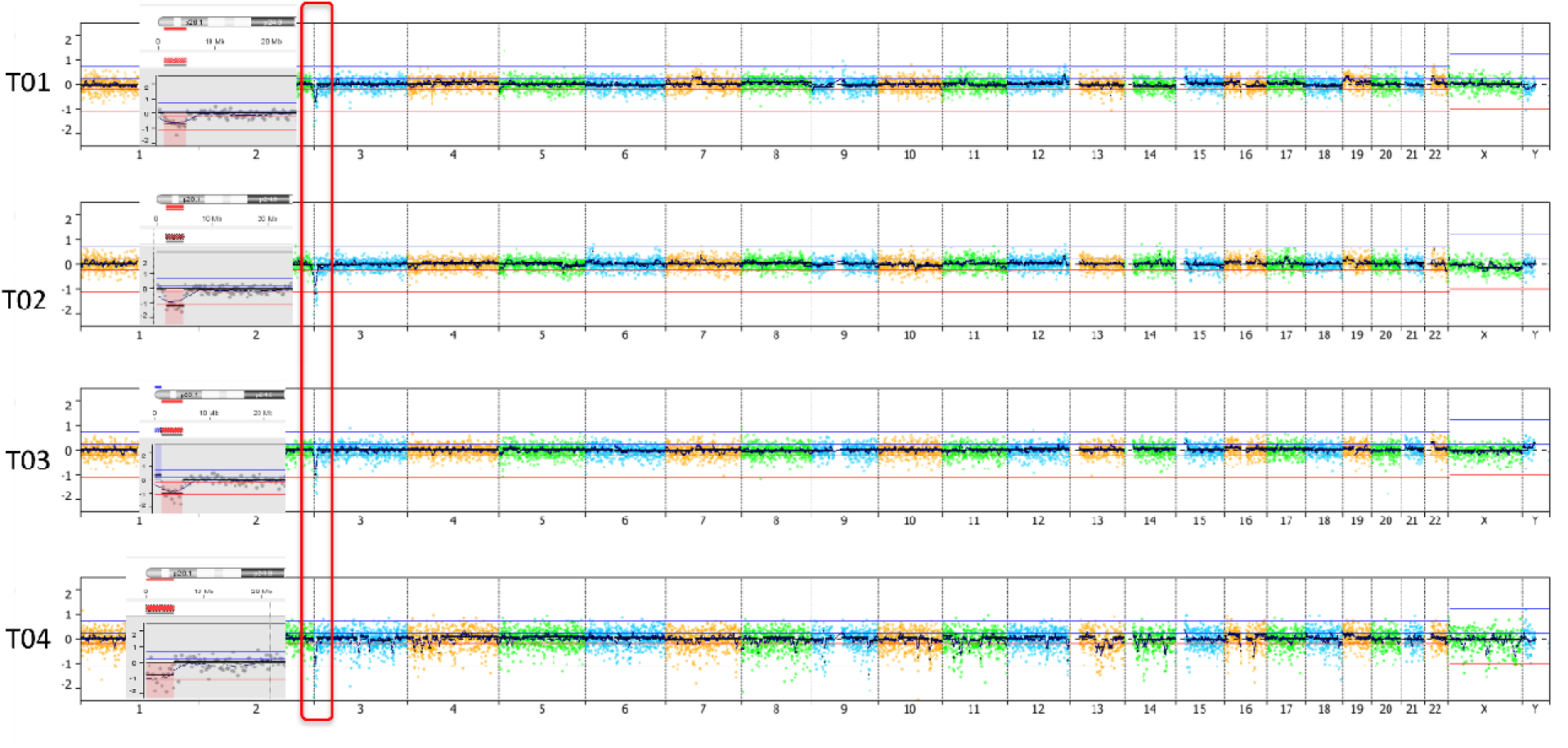
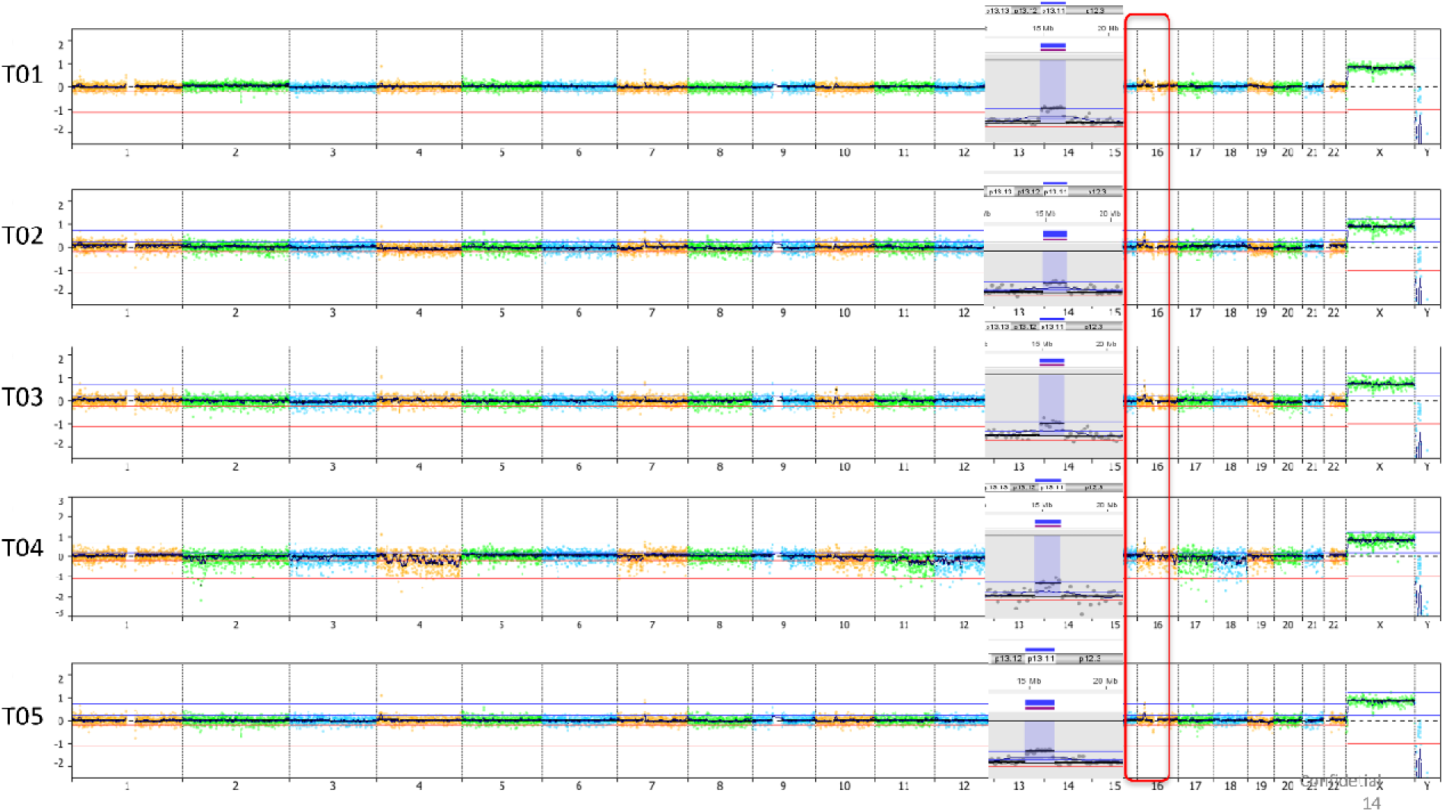
Plots from a deletion and a duplication in two different cases in the no-follow-up (NFU) series. Panel A shows del(3)(p26.3-p26.1 and panel B dup(16)(p13.11). The deleted and duplicated regions are in the red boxes on the genomic plots.

**FIGURE S4.**
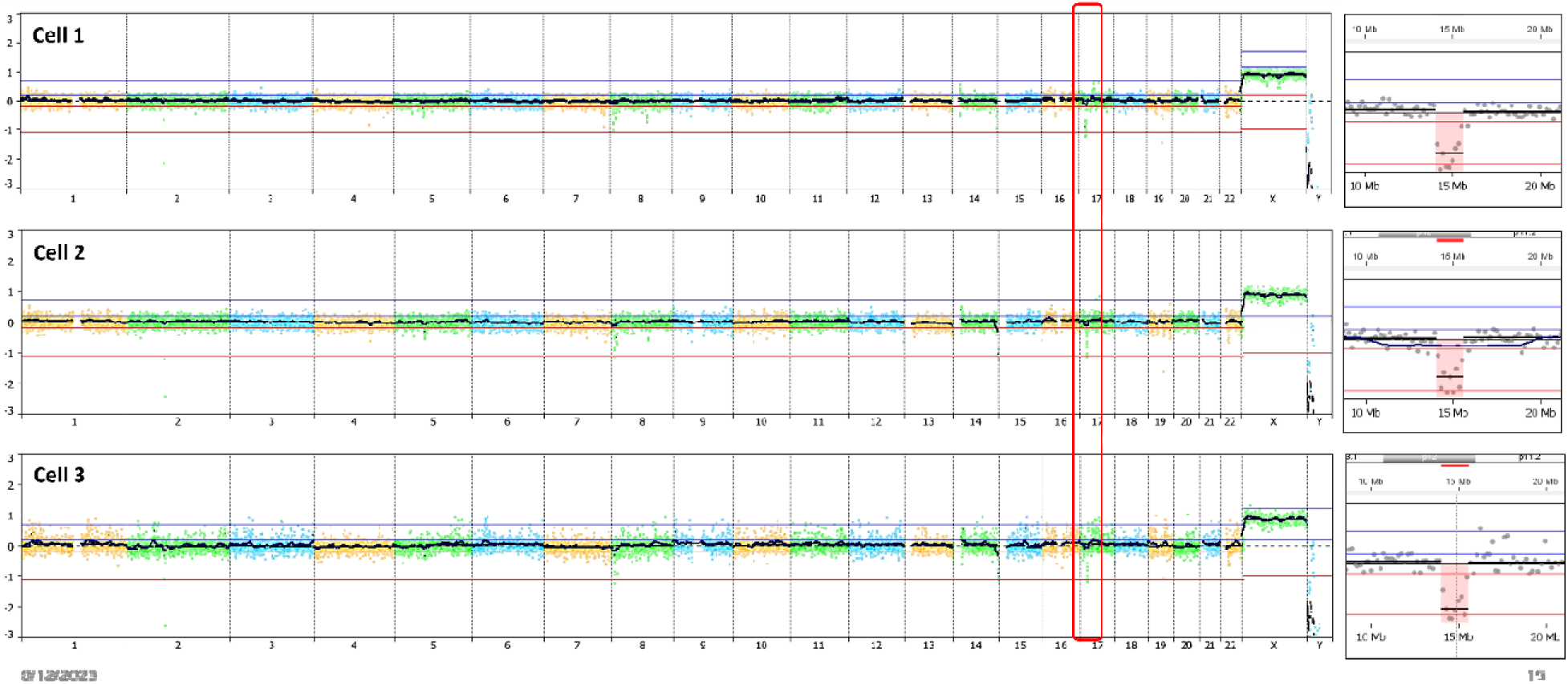
Data for three spike-in cells with del 17p12 including PMP22 gene. The deleted region is 1.5 Mb and is in the red box on the genomic plots. A small portion of chromosome 17 is shown in insets. Cells are male and plotted against a male control.

